# Retinal oxygen delivery and extraction in ophthalmologically healthy subjects with different blood pressure status

**DOI:** 10.1101/2021.09.20.21263850

**Authors:** Konstantinos Pappelis, Nomdo M. Jansonius

## Abstract

**Purpose:** To compare retinal oxygen delivery (DO_2_) and extraction (VO_2_) in ophthalmologically healthy subjects with different blood pressure (BP) status.

**Methods:** In this case-control study, we prospectively included 93 eyes of 93 subjects (age 50-65) from a large-scale population-based Dutch cohort (n=167,000) and allocated them to four groups (low BP, normal BP [controls], treated arterial hypertension [AHT], untreated AHT). We estimated vascular calibers from fundus images and fractal dimension (FD) from optical coherence tomography angiography scans. We combined calibers, FD, BP, and intraocular pressure measurements in a proxy of total retinal blood flow (RBF), using a validated Poiseuille-based model. We measured arterial and venous oxygen saturations (S_a_O_2_, S_v_O_2_) with a two-wavelength scanning laser ophthalmoscope. We calculated DO_2_ and VO_2_ from RBF, S_a_O_2_, and S_v_O_2_. We compared DO_2_ and VO_2_ between groups and investigated the DO_2_-VO_2_ association.

**Results:** DO_2_ and VO_2_ were different between groups (*P*=0.009, *P*=0.036, respectively). In post hoc analysis, the low BP group had lower DO_2_ than the untreated AHT group (*P*=4.9·10^-4^), while both the low BP group and the treated AHT group had lower VO_2_ than the untreated AHT group (*P*=0.021, *P*=0.034, respectively). There was a significant DO_2_-VO_2_ correlation (R_obs_=0.65, b_obs_=0.51, *P*=2.4·10^-12^). After correcting for shared measurement error, the slope was no longer significant (b_cor_=0.19, *P*=0.29), while the correlation coefficient could not be calculated.

**Conclusions:** DO_2_ and VO_2_ were altered in ophthalmologically healthy subjects with different BP status. Future studies could elucidate whether these changes can explain the increased risk of several ophthalmic pathologies in those subjects.

## Introduction

Blood pressure (BP) is implicated as a risk factor in the pathogenesis of several ophthalmic diseases, including leading causes of irreversible blindness, such as glaucoma, age-related macular degeneration (AMD), and diabetic retinopathy (DR).^1,2^ While each disease is characterized by distinct, complex pathogenetic mechanisms, the implication of blood flow and tissue oxygenation is considered to be a common denominator.^3^ However, to this day, our understanding of the influence of BP on the retinal oxygenation is largely incomplete.

It has long been known that the retina is a metabolically active tissue, thus prone to reduced oxygen (O_2_) supply due to hypoperfusion.^4,5^ Retinal blood flow (RBF) is mostly responsible for the O_2_ supply of the inner retinal layers through the superficial and deep capillary plexus.^6^ By diffusion, RBF also has a modest O_2_ contribution to the photoreceptors.^7^ BP is a major determinant of RBF, but, at the same time, its transient and chronic effects are dampened by tight autoregulatory mechanisms and vascular wall remodeling, which buffer the O_2_ volume delivered to the tissues (DO_2_).^8-10^ In addition, even when DO_2_ is eventually altered, human body tissues, including the retina, are still able to control the extraction of O_2_ volume (VO_2_) from the circulation, up to a certain extent.^11-13^ Consequently, it is difficult to a priori predict the effect of BP on retinal metabolism.

There is evidence that the concept of BP status could be more relevant to tissue oxygenation than BP alone, at least for certain ophthalmic diseases, such as glaucoma or ischemic optic neuropathy.^14-17^ Low BP (especially when presented as nocturnal dipping) can directly lead to hypoperfusion, while high BP can cause chronic damage to the endothelium, resulting in RBF dysregulation.^10,18^ Moreover, antihypertensive treatment may or may not fully protect the tissue from ischemic damage. This would depend on disease stage, on the individual contribution of certain medications, or even on low BP targets.^19-22^ The latter could bring the retinal vessels closer to their critical (lower) autoregulation limit, below which significant hypoperfusion may occur.^23-25^

Regardless, after the onset of any disease, it is almost impossible to disentangle the temporal relationship between perfusion deficits related to BP status and tissue apoptosis. Impaired oxygenation could simultaneously be the cause (reduced supply) and consequence (reduced demand) of cellular death. Therefore, in order to understand the involvement of BP in retinal disease, it is important, as a starting point, to establish how chronic BP status affects the oxygenation of the otherwise healthy retina.

The aim of this study was to compare absolute retinal O_2_ delivery and extraction in ophthalmologically healthy subjects with low BP, normal BP (controls), treated arterial hypertension (AHT), and untreated AHT. For this purpose, we used previously described approaches to combine static imaging-based modeling of absolute RBF with dual-wavelength retinal oximetry.

## Methods

### Study design and population

This is a cross-sectional, case-control study. We prospectively recruited subjects participating in Lifelines Biobank, an ongoing cohort study of the northern Netherlands (n≃167,000). The study comprised four groups, each one describing a distinct BP status: “low BP” (Group 1), “normal BP” (Group 2), i.e., controls, “treated AHT” (Group 3), and “untreated AHT” (Group 4). The group definitions were based on information from multiple (at least two) previous visits. The exact definitions and rationale have been extensively described in our recent study on the same population.^25^ In short, we required both the systolic and diastolic BP (SBP, DBP) of subjects belonging to Groups 1 and 4 to consistently belong to the lowest and highest deciles of the Lifelines distribution, respectively. We also required both SBP and DBP in Group 2 to fall no more than 1 standard deviation (SD) away from their means. Last, for subjects in Group 3, we required uninterrupted use of antihypertensive medication for at least the past year. Invitations were sent to participants between 50 and 65 years old satisfying these BP criteria. Subjects that responded to our invitation underwent further screening for ophthalmic conditions and a general medical history interview. For each study group, achievement of predetermined power levels (or lack of participant availability) was considered as the end of the recruitment.

We excluded participants with best-corrected visual acuity less than 0.8 (20/25), spherical refractive error larger than +3 D or -3 D, cylinder exceeding 2 D, IOP higher than 21 mmHg (non-contact tonometer Tonoref II, Nidek, Aichi, Japan), reproducibly abnormal visual field test locations (Frequency Doubling Technology [C20-1 screening mode], Carl Zeiss, Jena, Germany), family history of glaucoma, and any ophthalmic pathology, including history of previous ophthalmic surgery. Absence of ophthalmic disease was confirmed with the subsequent imaging sessions (see below). We also excluded participants with diabetes, cardiovascular disease (except for AHT in Groups 3 and 4), hematologic disease, and lung disease. We did not exclude smokers, but, since early chronic obstructive pulmonary disease cannot be completely ruled out in these participants, we recorded any previous or current regular smoking.

All participants provided written informed consent. The ethics board of the University Medical Center Groningen approved the study protocol (#NL61508.042.17). The study followed the tenets of the Declaration of Helsinki.

### Retinal blood flow measurements

In total, 105 participants satisfying the BP definitions and screening criteria qualified for the subsequent imaging session. Before the start of the imaging session, we performed standard on-site BP measurements. The collected imaging data relevant to this study were, in short: ONH-centered fundus images, 6×6 mm OCTA macula scans, and retinal images obtained with a scanning laser ophthalmoscope (SLO). The details behind fundus imaging and OCTA scans can be found in our related study on the same population, while the details behind the SLO scanning are provided in the next subsection.^25^ For the imaging session, we selected one eye per participant. If both eyes satisfied the ophthalmic inclusion criteria, this selection was random.

RBF estimations were based on a static imaging model-based approach. This protocol has been shown to have very good agreement with in vivo RBF measurements of the human retina, assessed by Laser Speckle Flowgraphy (LSFG).^24^ Hereunder, we provide an outline of the procedure. After data collection, we first estimated the central retinal artery and vein equivalents (CRAE, CRVE), i.e., vascular calibers, from fundus images and the microvascular branching complexity (fractal dimension; FD) from en face optical coherence tomography angiography (OCTA) scans, using standard methods.^26-28^ We then calculated total retinal vascular resistance (RVR) for each subject, by combining these measurements with blood viscosity in a Poiseuille-based fractal branching model.^24,29^ Lastly, we calculated RBF from RVR and refined estimates of retinal perfusion pressure (RPP):

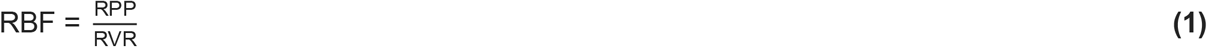

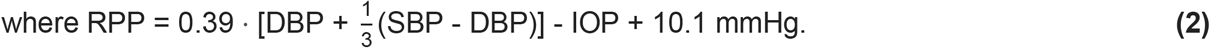

OCT and OCTA scans with image quality less than 7/10, segmentation errors, or artifacts were excluded.^30^ This resulted in the exclusion of 9 subjects. All subjects had high-quality fundus images.

### Oxygen saturation measurements

An SLO (Optomap 200Tx, Optos PLC, Dunfermline, United Kingdom) was used to measure arterial and venous O_2_ saturations (S_a_O_2_, S_v_O_2_), by means of a commonly used dual-wavelength technique.^31,32^ The device simultaneously acquires two retinal images, one at a wavelength of 532 nm (“oxygen-insensitive”; Figure 1A) and the other at a wavelength of 633 nm (“oxygen-sensitive”; Figure 1B). Three images per eye were obtained with the ResMax option (approximately 60°), at the same laser intensity, and with the gain set at medium iris pigmentation. Three more subjects were excluded, due to persistent blinking artifacts.

**Figure 1.**
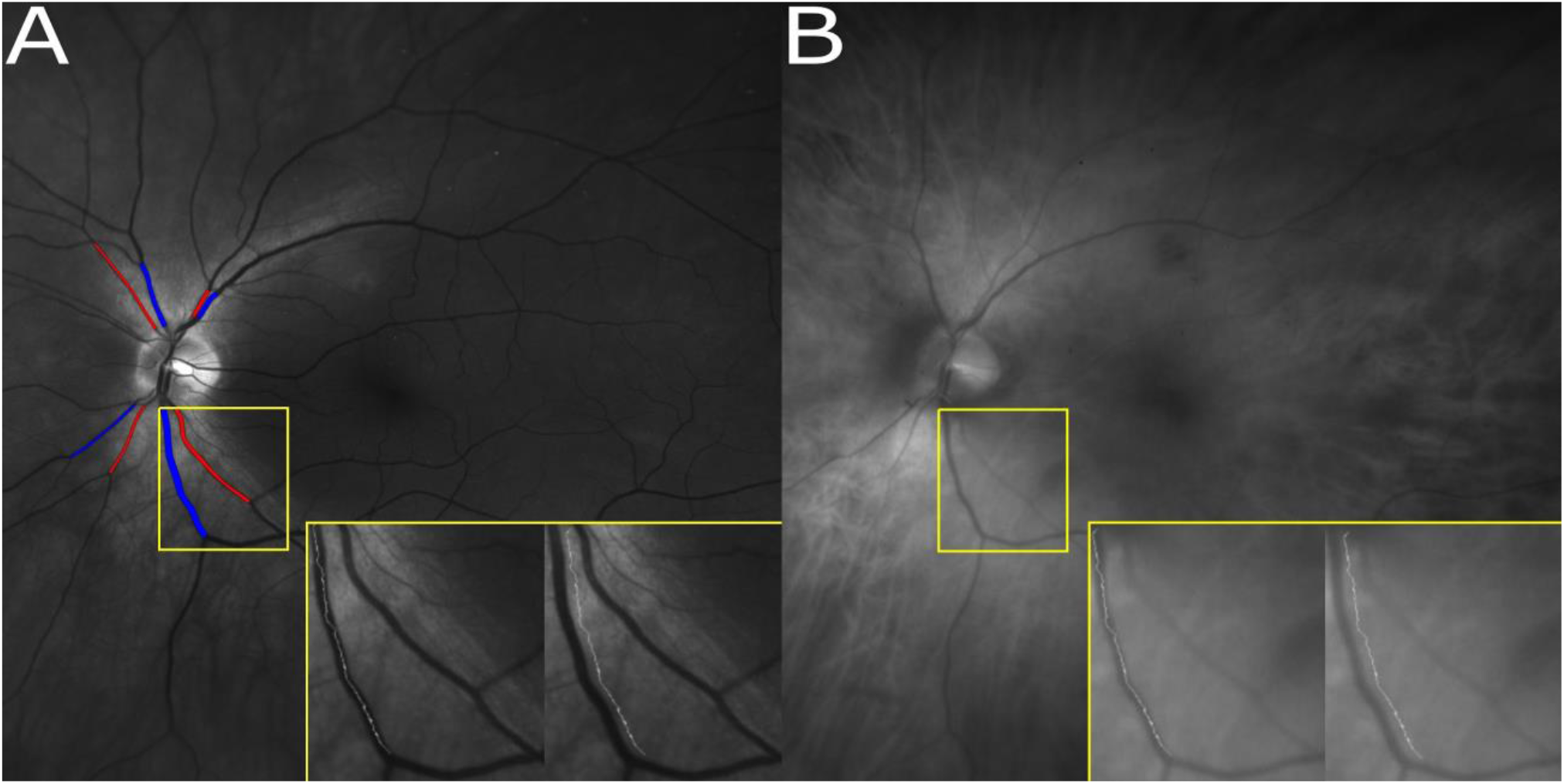
A) Scanning laser ophthalmoscope (SLO) image at 532 nm. At this isosbestic (“oxygen-insensitive”) wavelength, arteries and veins appear similar, in terms of optical density. Measured arterial and venous segments are marked in red and blue, respectively. The path of minimal intensity inside the vessel and a parallel path outside the vessel are shown for the inferotemporal vein. B) SLO image at 633 nm. At this non-isosbestic (“oxygen-sensitive”) wavelength, arteries appear brighter than veins, due to higher O_2_ content.

The optical density (OD) of a vessel at a given wavelength is defined as:

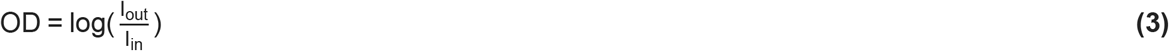

where I_out_ is taken equal to the average grayscale intensity over a measurement area outside the vessel and I_in_ is taken equal to the average grayscale intensity inside the vessel.

The optical density ratio (ODR) of a vessel at two given wavelengths (in this case, 633 nm and 532 nm) is defined as:

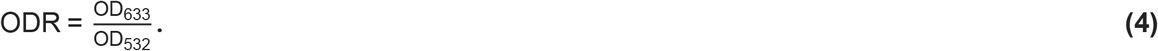

By using the Beer-Lambert law, it can be shown that, under ideal conditions:

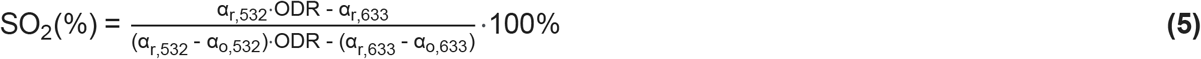

where α_r_ is the absorption coefficient of reduced hemoglobin (Hb) and α_o_ is the absorption coefficient of oxyhemoglobin (HbO_2_).

Since 532 nm is an almost isosbestic wavelength (α_r,532_≃α_o,532_) and ODR is small, it is commonly assumed (and experimentally verified) that SO_2_ falls linearly with increasing ODR, that is:

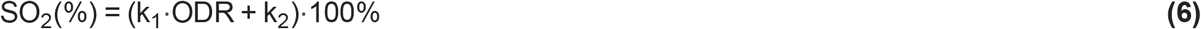

where k_1_, k_2_ are constants determined after calibration.^31,33,34^

In order to obtain saturation values for the central retinal artery and vein (CRA, CRV), we first need to measure the saturation of their visible branches. We selected the largest artery and vein of each quadrant (superotemporal [ST], inferotemporal [IT], superonasal [SN], inferonasal [IN]) and ignored smaller vessels, since this approach has been found to minimally affect estimations.^35^ To minimize O_2_ diffusion losses, measurements were taken close to the border of the optic disc, according to a previously described, semi-automatic protocol.^34^ In short, the path of minimal intensity (thus, avoiding the vessel light reflex) between the starting point of the vessel and the first major branching was automatically traced. A parallel path outside the vessel, at a fixed distance of 30 pixels was also automatically traced (Figure 1). ODR was subsequently calculated, according to Eqs. 3 and 4, with the image grayscale value as the standard proxy for intensity.^33^ For each vessel, we recorded the median of three measurements. Feasibility, repeatability, and reproducibility of this approach have been previously established, also for an SLO.^36-40^

Obtained ODR values have to be corrected for the artifactual influence of factors other than O_2_ saturation, mostly related to magnification errors, heterogeneous light absorption, and photon backscattering, before they can be used to calculate SO_2_ via Eq. 6.^31,33,41^ For each vessel segment, we implemented linear compensations by means of a backward regression model, with ODR serving as the dependent variable and the independent variables being potential confounders, i.e., laterality, optic disc area, spherical error, cylinder, vessel diameter, and quadrant pigmentation index (PI). Only the significant variables of the reduced (final) model were subsequently used to construct the following correction formula:

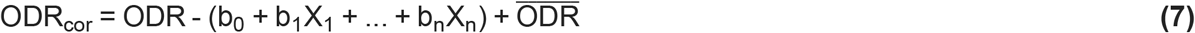

where ODR_cor_ is the corrected ODR, b_i_ are the regression coefficients, X_i_ are the confounders that remained in the reduced model, and 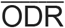 is the average ODR of the study population. ODR_cor_ was evaluated for each quadrant separately.

PI was calculated based on extravascular reflection, as follows:

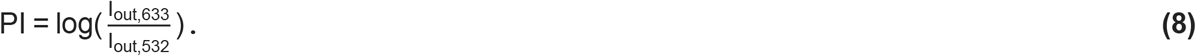

Since the increase in light absorption with increasing melanin is more pronounced at longer wavelengths, lower PI values indicate increased pigmentation.^42^

O_2_ saturation in the CRA can be calculated as the average measured saturation in the four major arterial arcades:

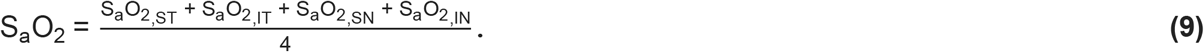

O_2_ saturation in the CRV can be calculated as the average measured saturation in the four major venous arcades, weighted by the relative flow contribution of each arcade. The weigh factor equals a power of the radius of the relevant venular segment;^43^ the Poiseuille-based model used for absolute RBF estimations assumes that the power is equal to FD+1.15, where FD is the 2D-fractal dimension measured by OCTA (see *Retinal blood flow measurements*) and 1.15 is a branch length coefficient.^24,29,44^ Therefore:

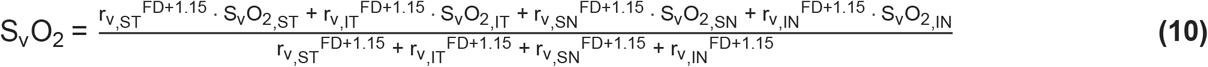

where r_v_ denotes the radius of the relevant venular segment.

Using Eqs. 6, 7, 9, and 10, we can express S_a_O_2_ and S_v_O_2_ as a function of the regression coefficients b_i_ and the constants k_1_ and k_2_. As described above, the b_i_ can be calculated from a standard linear fitting. Now, to determine k_1_ and k_2_, we need two distinct calibration values. The average S_a_O_2_ and S_v_O_2_ values for healthy eyes reported by Schweitzer *et al*. (92.2% and 57.9%, respectively) are the values most frequently used for calibration.^32^ With these values, the calibration constants were k_1_ = -2.46 and k_2_ = 1.26. Calibration is unrestrictive and allows saturation measurements to exceed 100%, due to variability.^37,45^

### Total retinal oxygen delivery and extraction

We can now estimate the outcome variables, DO_2_ and VO_2_, from the Fick principle, as demonstrated by Werkmeister *et al*.^46^:

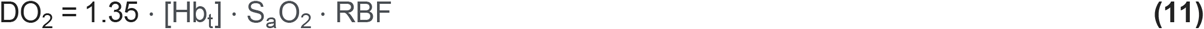

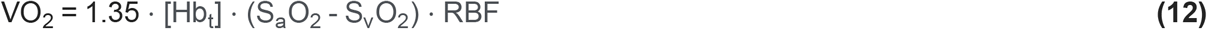

where [Hb_t_] is the total hemoglobin concentration.

In both formulas, we omitted a term for unbound O_2_ content, since it is smaller than bound O_2_ content by more than two orders of magnitude.^47^ We did not obtain blood samples and we therefore used the average [Hb_t_] values reported in the Lifelines Biobank, stratified by age, sex, and blood pressure status. For this reason, we also conducted a sensitivity analysis, repeatedly replacing the average [Hb_t_] values with random values taken from identical distributions. Lastly, the oxygen extraction fraction (OEF) was defined as:

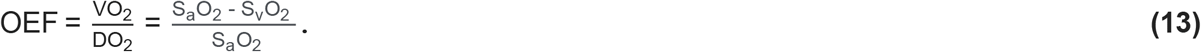

### Statistical analysis

We compared general characteristics and outcome variables (DO_2_ and VO_2_) between the four groups, by means of ANOVA models. Potential confounding factors from the population general characteristics were included as covariates in the outcome variable models. In post hoc analysis, we used the Tukey HSD correction to account for multiple comparisons. Whenever ANOVA assumptions were not met, we used Welch’s ANOVA or non-parametric tests.

To examine the overall association between DO_2_ and VO_2_, we used linear regression analysis. The observed correlation of these two variables is inflated, due to shared measurement error, stemming from the mathematical coupling of these variables. This phenomenon is commonly reported in other systems.^13^ We used the method described by Stratton *et al*. to calculate the corrected regression coefficient.^48^ In short, we calculated a reliability coefficient r_D_:

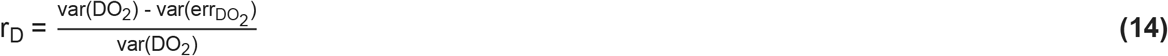

where var(DO_2_) denotes the variance of DO_2_ and var(err_DO2_) denotes the variance attributable to measurement error.

The variance attributable to measurement error, var(err_DO2_), was calculated from the error variances of RBF and S_a_O_2_, used in the calculation of DO_2_ (Eq. 11). As described by Stratton *et al*., these error variances need to be derived from calibration.^48^ In the absence of a gold calibration standard, the error variance of RBF was approximated from a previous experiment in an independent population, from the residual variance of RBF fitted on in vivo LSFG measurements.^24^ Since the physiological variation of S_a_O_2_ in healthy individuals is expected to be minimal, the error variance of S_a_O_2_ was set as equal to the observed variance of S_a_O_2_. The corrected slope (b_cor_) for VO_2_ as a function of DO_2_ can then be calculated as:

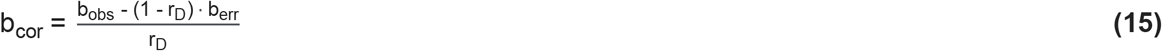

where b_obs_ is the observed slope and b_err_ the slope of measurement errors, which accounts for the covariance of the error in DO_2_ and VO_2_. We will not demonstrate here the detailed mathematical calculations of var(err_DO2_) and b_err_, as they are extensively provided in the aforementioned paper by Stratton *et al*.^48^

Henceforth, all normally distributed variables are described with the mean and SD. Variables with a skewed distribution are described with the median and interquartile range (IQR). All analyses were performed using R (version 3.3.3; R Foundation for Statistical Computing, Vienna, Austria) and SPSS (version 26; IBM Corp., Armonk, NY). A *P* value of 0.05 or less was considered statistically significant.

## Results

### General characteristics

We excluded 12 out of 105 participants (3 from the “low BP” group, 3 from the “control” group, 4 from the “treated AHT” group, and 2 from the “untreated AHT” group), due to artifacts or insufficient image quality at any stage of the imaging session. This resulted in a total of 93 eyes from 93 participants being included in the analysis. Their general characteristics are summarized in Table 1. The BP measurements in this table represent the actual on-site measurements preceding the scanning; the highly-significant *P* values confirm the robustness of the inclusion procedure. Groups differed furthermore significantly in sex and BMI, which is expected due to the prevalence of low BP in females and high BMI among hypertensives, also verified in the Lifelines cohort.^49,50^ Therefore, we adjusted subsequent analyses (see next subsection) for sex and BMI. We additionally adjusted DO_2_ and VO_2_ analyses for smoking status (despite not being significantly different among groups), since it could affect O_2_ metrics.

**Table 1.** Characteristics of the study population.

|  | **Group 1**<br>**(Low BP)** | **Group 2**<br>**(Normal BP)** | **Group 3**<br>**(Treated AHT)** | **Group 4**<br>**(Untreated AHT)** | ***P* value** |
| --- | --- | --- | --- | --- | --- |
| **Group size (N)** | 30 | 20 | 26 | 17 |  |
| **Age; years**<br>**[median (IQR)]** | 56.0<br>(51.0 to 59.3) | 53.5<br>(51.3 to 60.5) | 55.5<br>(52.8 to 61.0) | 58.0<br>(53.0 to 61.0) | 0.74 |
| **Sex; % female** | 93.3 | 50.0 | 42.3 | 47.1 | **2.2·10<sup>-4</sup>** |
| **SBP; mmHg**<br>**[mean (SD)]** | 107 (9) | 125 (5) | 142 (18) | 159 (23) | **2.1·10<sup>-14</sup>** |
| **DBP; mmHg**<br>[mean (SD)] | 66 (6) | 79 (6) | 86 (11) | 99 (8) | **4.1·10<sup>-23</sup>** |
| **BMI; kg·m<sup>-2</sup>**<br>[median (IQR)] | 22.4<br>(21.2 to 24.3) | 23.4<br>(22.2 to 26.7) | 26.9<br>(24.7 to 29.8) | 27.4<br>(24.3 to 28.5) | **5.0·10<sup>-6</sup>** |
| **Smoking; % yes** | 23.3 | 35.0 | 30.8 | 41.2 | 0.62 |
| **IOP; mmHg**<br>[mean (SD)] | 14.0 (3.0) | 13.4 (3.1) | 14.3 (3.0) | 14.5 (3.8) | 0.72 |
| **SEQ; D**<br>[mean (SD)] | -0.11 (1.44) | +0.19 (1.67) | -0.23 (1.55) | -0.65 (1.58) | 0.45 |
| **ONH area; mm<sup>2</sup>**<br>[median (IQR)] | 1.89<br>(1.68 to 2.24) | 1.97<br>(1.71 to 2.20) | 1.94<br>(1.72 to 2.31) | 1.98<br>(1.78 to 2.15) | 0.81 |
| BP, blood pressure; AHT, arterial hypertension; SD, standard deviation; SBP, systolic blood pressure; DBP, diastolic blood pressure; BMI, body mass index; IOP, intraocular pressure; SEQ, spherical equivalent; ONH, optic nerve head. |  |  |  |  |  |

Supplementary Table S1 displays the significant variables of the reduced linear models used for the ODR correction. Corrected ODR values per retinal quadrant and the global averages are provided in Supplementary Table S2. In general, aside from O_2_ content, ODR values were also influenced by measurement location (quadrant), pigmentation, refractive errors, and, in some cases, eye laterality (see Discussion).

### Associations with BP status

Table 2 summarizes the measured components used in the estimation of DO_2_ and VO_2_, stratified by BP status. Groups were similar in terms of S_a_O_2_ and S_v_O_2_, but differed in terms of RBF, in univariable analysis. Sex, BMI, and smoking history did not confound this relationship when included as covariates, nor did they after omitting each one of the four groups from the analysis (*P*>>0.05 in all cases). In post hoc analysis, after adjusting for multiple comparisons, the low BP group had a significantly lower mean RBF than the untreated AHT group (*P*=0.032).

**Table 2.** Group summaries for vascular and oximetry measurements.

|  | **Group 1**<br>**(Low BP)** | **Group 2**<br>**(Normal BP)** | **Group 3**<br>**(Treated AHT)** | **Group 4**<br>**(Untreated AHT)** | **P value** |
| --- | --- | --- | --- | --- | --- |
| **Total RBF; $\mu\text{L}\cdot\text{min}^{-1}$**<br>**[mean (SD)]** | 40.7 (6.0) | 45.7 (9.6) | 45.1 (8.6) | 47.6 (8.3) | **0.028** |
| **RPP; mmHg**<br>**[mean (SD)]** | 27.2 (2.8) | 33.5 (3.9) | 36.6 (4.7) | 41.2 (5.1) | **$1.6 \cdot 10^{-19}$** |
| **CRAE; $\mu\text{m}$**<br>**[mean (SD)]** | 172 (12) | 162 (12) | 154 (13) | 148 (7) | **$7.0 \cdot 10^{-10}$** |
| **CRVE; $\mu\text{m}$**<br>**[mean (SD)]** | 228 (18) | 229 (16) | 229 (18) | 223 (12) | 0.57 |
| **OCTA FD**<br>**[mean (SD)]** | 1.625 (0.005) | 1.626 (0.007) | 1.624<br>(0.006) | 1.626<br>(0.006) | 0.67 |
| **[Hb<sub>t</sub>]; $\text{g}\cdot\text{dL}^{-1}$ [mean (SD)]**<br>**(population-based data)** | 13.6 (0.8) | 14.3 (0.8) | 14.2 (0.9) | 14.7 (0.8) | **$5.3 \cdot 10^{-307}$** |
| **S<sub>a</sub>O<sub>2</sub> (%) [mean (SD)]** | 94.7 (12.8) | 88.6 (12.4) | 89.3 (11.2) | 96.3 (12.7) | 0.12 |
| **S<sub>v</sub>O<sub>2</sub> (%) [mean (SD)]** | 59.1 (14.2) | 53.2 (13.6) | 60.2 (14.5) | 56.3 (14.8) | 0.36 |

BP, blood pressure; AHT, arterial hypertension; RBF, retinal blood flow; SD, standard deviation; RPP, retinal perfusion pressure; CRAE, central retinal artery equivalent; CRVE, central retinal vein equivalent; OCTA, optical coherence tomography angiography; FD, fractal dimension; [Hb<sub>t</sub>], total hemoglobin concentration; S<sub>a</sub>O<sub>2</sub>, arterial oxygen saturation; S<sub>v</sub>O<sub>2</sub>, venous oxygen saturation.
\* Data and comparison taken from Lifelines (n=28,888).

Figure 2 shows the final outcome variables, DO_2_ and VO_2_, stratified by BP status. Both variables were significantly different between groups (*P*=0.009 and *P*=0.036, respectively) in univariable analysis and the significance of this relationship was largely unaffected when [Hb_t_] values were repeatedly replaced with random values taken from the same distribution (DO_2_: *P*[average]=0.008, *P*[95% of repetitions]=0.002-0.028; VO_2_: *P*[average]=0.025, *P*[95% of repetitions]=0.014-0.044). The significance of this relationship was also unaffected when sex, BMI, and smoking history were included as covariates (Table 3). All three covariates had additional, independent effects, which are summarized in Table 3 (reported as coefficients from the equivalent generalized linear models). In post hoc analysis, after adjusting for multiple comparisons, the low BP group had a significantly lower estimated marginal mean DO_2_ than the untreated AHT group (*P*=4.9·10^-4^), while both the low BP group and the treated AHT group had a significantly lower estimated marginal mean VO_2_ than the untreated AHT group (*P*=0.021 and *P*=0.034, respectively).

**Figure 2.**
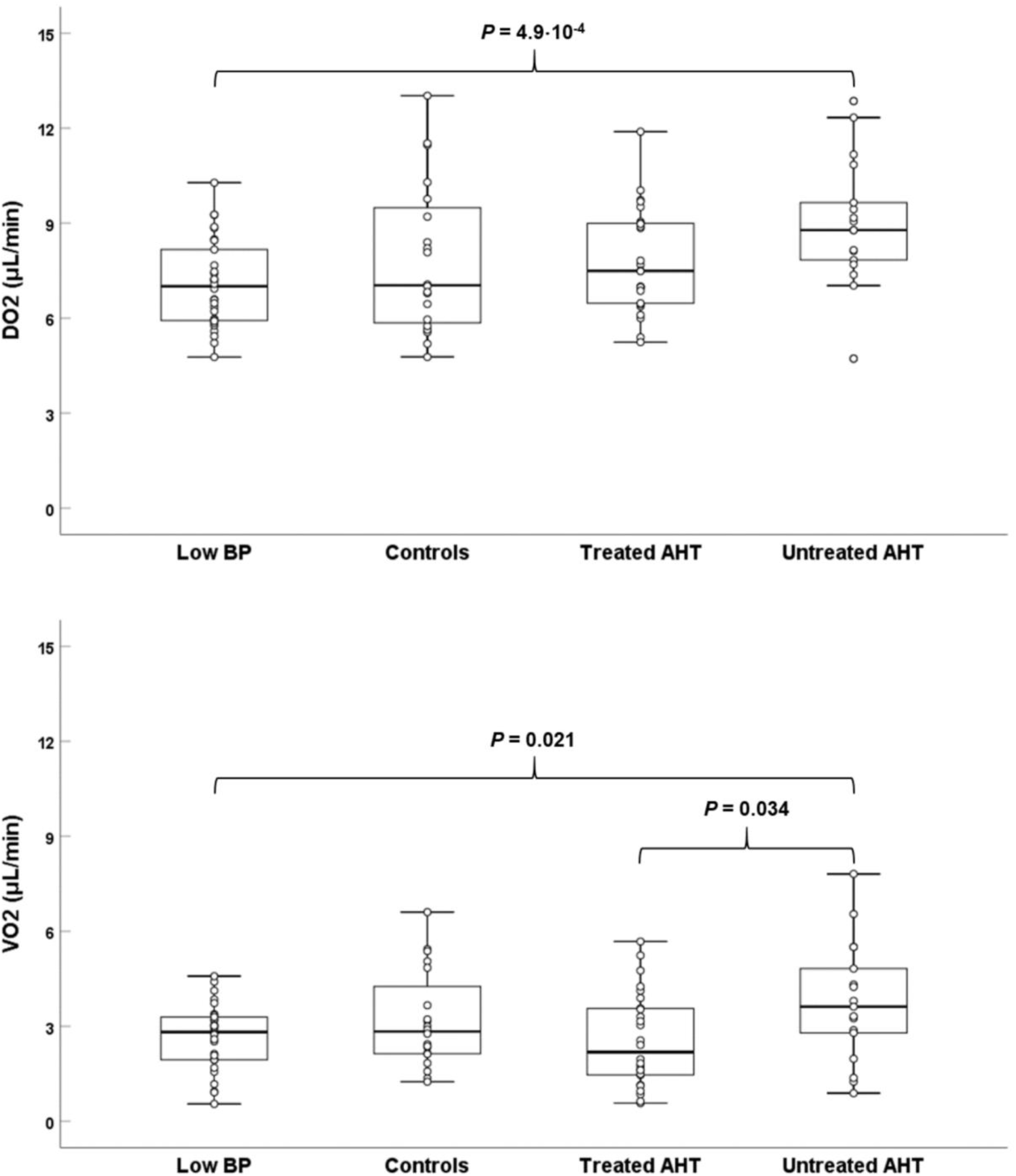
Total retinal oxygen delivery (DO_2_) and extraction (VO_2_) as a function of blood pressure (BP) status. DO_2_ is higher in subjects with untreated arterial hypertension (AHT), compared to the low BP group. VO_2_ is higher in subjects with untreated AHT, compared to both the low BP and the treated AHT group.

**Table 3.** Factors associated with DO_2_ and VO_2_ in multivariable analysis

| | $DO_2$ ($\mu\text{L} \cdot \text{min}^{-1}$) | | $VO_2$ ($\mu\text{L} \cdot \text{min}^{-1}$) | |
| --- | --- | --- | --- | --- |
|  | Coefficient | P value | Coefficient | P value |
| **BP status;**<br>**ref.: Untreated AHT** |  |  |  |  |
| Low BP | -2.4 | **$2.2 \cdot 10^{-5}$** | -1.4 | **0.002** |
| Normal BP | -1.5 | 0.013 | -0.9 | 0.042 |
| Treated AHT | -1.1 | 0.041 | -1.2 | **0.003** |
| **BMI; $\text{kg} \cdot \text{m}^{-2}$** | -0.14 | **0.027** | -0.13 | **0.006** |
| **Sex; female=1** | NS | NS | -0.7 | **0.038** |
| **Smoking; yes=1** | *NS* | *NS* | -0.7 | **0.017** |
| DO<sub>2</sub>, total retinal oxygen delivery; VO<sub>2</sub>, total retinal oxygen extraction; BP, blood pressure; AHT, arterial hypertension; BMI, body mass index; *NS*, not significant.<br>**For the BP status, $P=0.008$ is taken as the adjusted threshold of significance.** |  |  |  |  |

Mean (SD) OEF was 0.37 (0.15) and was similar between groups (*P*=0.20). In multivariable analysis, smoking was associated with a decrease in the OEF (b=-0.07, *P*=0.039).

### DO_2_-VO_2_ relationship

The pooled (n=93) association of DO_2_ and VO_2_ is plotted in Figure 3. Initially, there was a significant positive correlation between the two variables (R_obs_=0.65, b_obs_=0.51, *P*=2.4·10^-12^). However, after correcting for shared measurement error due to mathematical coupling, the true slope was flatter and no longer statistically significant (b_cor_=0.19, *P*=0.29), while the true correlation coefficient could not be calculated.^48^

**Figure 3.**
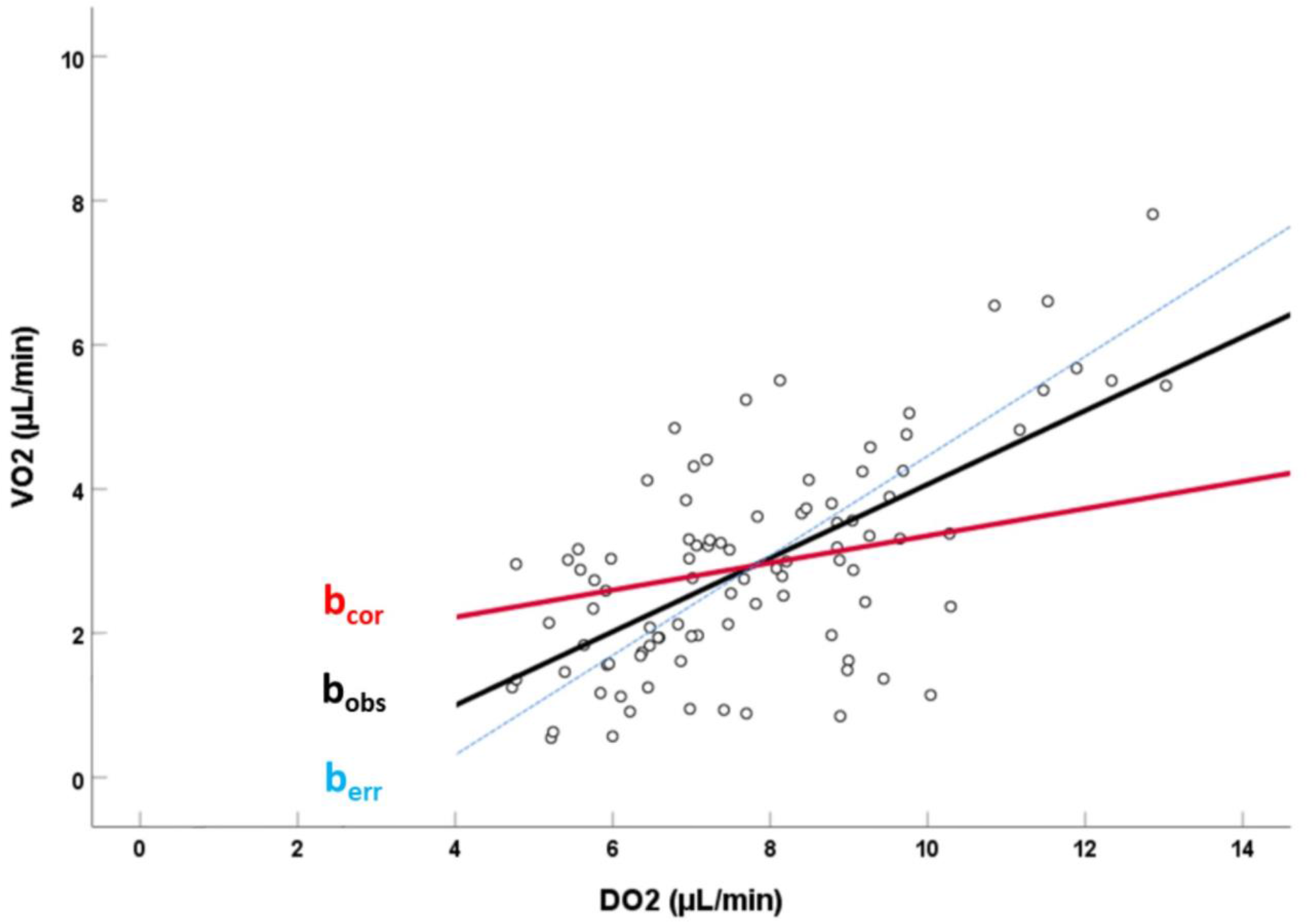
Pooled (n=93) data for total retinal oxygen extraction (VO_2_) plotted as a function of total retinal oxygen delivery (DO_2_). The observed slope of the linear relationship (continuous black line) is denoted by b_obs_. The component of the slope (dashed blue line) introduced purely via shared measurement error, due to mathematical coupling of the two variables, is denoted by b_err_. The true, corrected slope of the relationship (continuous red line), i.e., adjusted for shared measurement error, is denoted by b_cor_. The small, non-significant slope of the true relationship indicates little interdependency of resting DO_2_ and VO_2_.

## Discussion

In this study, we showed that BP status was associated with altered total retinal oxygen delivery (DO_2_) and extraction (VO_2_) at rest, in ophthalmologically healthy subjects. Specifically, we reported an increase in DO_2_ and VO_2_ with increasing BP, which was more prominent at the tails of the BP distribution. In addition, VO_2_ was also higher in subjects with untreated AHT than in subjects with treated AHT. The true association between DO_2_ and VO_2_ was, at best, weak, suggesting these variables are mostly independent at rest.

### Oxygen delivery (DO_2_)

Although average RPP increased by 51% on the way from the low BP group to the untreated hypertensives (Table 3), average RBF only increased by 17% (Table 3) and average DO_2_ by 27% (Table 3, Figure 2). As a result, differences in RBF and DO_2_ were noted as significant only between the groups representing the tails of the BP distribution. This observation is not surprising, as it is consistent with the general concept of RBF autoregulation and with the subsequent structural remodeling of blood vessels, following chronic BP elevation.^8,10,51^ Experimental studies and mathematical models have shown an increase in the tissue oxygen partial pressure (PO_2_) with transient BP manipulations.^52,53^ Our results suggest that chronic low or high, but still physiological, resting BP values can also have a subtle effect on DO_2_. The increase in DO_2_ (which is the product of RBF and O_2_ content) with increasing BP was larger than that of RBF alone. Since S_a_O_2_ was similar between groups (Table 3), this is mostly due to the increasing [Hb_t_] with increasing BP (Table 3). This association is known from other population studies.^54,55^

Higher BMI was found to be independently associated with reduced DO_2_, but not RBF, which likely reflects the documented effects of obesity on lung function and pulmonary gas exchange.^56^

### Oxygen extraction (VO_2_)

A significantly higher VO_2_ was also observed in untreated hypertensives (Table 3, Figure 2), relatively to subjects with low BP (+41%) and treated hypertensives (+46%). Again, no group was significantly different from controls, after adjusting for multiple comparisons (Table 3). The smaller effect size, in combination with the high variability that is present in our estimates, seems the most likely explanation for this phenomenon. Indeed, while our average estimates for both DO_2_ and VO_2_ are similar to previously published values (and almost identical when the same calibration values are used), the variability we report is higher.^46,47^ This was due to the larger error introduced by the SLO-based retinal oximetry (which is consistently present in all studies using the SLO), compared to the error yielded by tailored fundus cameras.^37,39,40^

The reported alterations in VO_2_ can have a number of pathophysiological explanations. We have recently shown that, in the same population, the low BP group and the treated hypertensives were characterized by subtle thinning of the ganglion cell-inner plexiform layer and the macular and peripapillary retinal nerve fiber layer.^25^ While we do not know for sure if these insults are primary or secondary to reduced DO_2_, lower VO_2_ in these groups could, at least partially, suggest a current state of reduced O_2_ demand. However, thinning of the inner retina was also present (albeit less pronounced) in the untreated hypertensive group, which contradicts the higher average VO_2_ reported for these subjects. In this regard, in the next paragraphs we speculate about potential mechanisms that could explain this finding.

Firstly, it is estimated that about 15% of the O_2_ extracted from the retinal circulation is not consumed by the inner retina, but actually complements the choroidal O_2_ supply of the photoreceptors in the outer retina. As a result, any condition that compromises the choroidal supply could lead to a compensatory increase in retinal VO_2_.^7^ This VO_2_ increase has already been observed in mice with early diabetes, while it has also been reported that deficits of the deep retinal capillary plexus in human diabetics are associated with photoreceptor loss.^57,58^ This could also apply to AHT, since it has been shown that AHT, especially when poorly controlled, has a detrimental effect on the choriocapillaris.^59^ The magnitude of this effect is unknown, but it should be able to counteract opposing forces that would tend to reduce VO_2_, at least theoretically, such as reduced mean circulation time, shunting of flow, and deep capillary rarefaction.^60,61^

Secondly, experimental evidence and mathematical models support that, in the presence of increased O_2_ availability, the O_2_ consumption of the inner retina (especially that of the inner and outer plexiform layers) increases, a mechanism that helps to keep inner retinal O_2_ levels relatively stable.^6,52,62,63^ This could pertain to subjects with untreated, chronic AHT. Indeed, while hyperoxic conditions such as 100% oxygen breathing achieve control O_2_ levels mostly by inducing RBF reduction through vasoconstriction, this is obviously not possible in conditions where RBF is a priori increased, despite vessels being already constricted.^64^ Increased resting consumption in these subjects could, at least partially, explain increased VO_2_.

That said, the true correlation analysis for DO_2_-VO_2_ indicates that this increase in resting O_2_ consumption with increased resting delivery cannot be too pronounced, if any. Indeed, while mitochondrial oxidative phosphorylation could act as a sensor of O_2_ tension, it also has a saturation ceiling.^65^ It is not impossible, nevertheless, that a metabolic transition to a more oxidative state, with reduced glycolytic activity, occurs.^66^ Other factors could also play a role in determining the DO_2_-VO_2_ slope. With constant consumption, a rise in BP would initially cause an increase in RBF (and DO_2_), which would be compensated by autoregulation. Now, with constant BP, an increase in consumption would cause a rise in local CO_2_, inducing vasodilation, thus causing an increase in DO_2_. The gain of this feedback loop at equilibrium is possibly a major determinant of this slope and is, therefore, likely close to zero.

Higher BMI was, again, independently associated with reduced VO_2_, possibly pointing towards reduced retinal function due to oxidative stress and inflammation.^67^ VO_2_ was lower in females than males, on which conflicting evidence exists, and could reflect the lower systemic basal metabolic rate.^47,68^ Smoking was also associated with reduced VO_2_ and could be related to the reported reduced O_2_ uptake present in other systems. Lastly, although the average, resting OEF was consistent with previous reports, we found that it was lower in smokers.^11^ Studies with a tailored design could shed more light on these secondary observations.

### Study strengths and limitations

To our knowledge, this is the first study describing alterations in DO_2_ and VO_2_ with different chronic BP status. The strict selection process that we implemented within the setting of a large-scale cohort (Lifelines Biobank) enabled us to investigate the true tails of the BP distribution. Another novelty of our study is the fact that we describe the true resting DO_2_-VO_2_ relationship in the retina, which avoids overestimations originating from mathematical coupling. However, there are several limitations that should be considered.

Firstly, the calculation of outcome variables (DO_2_ and VO_2_) is liable to propagation of error associated with their measured components. There is currently no gold standard way to measure these components in the clinic and there are still unresolved technical considerations with regards to acquisition and quantification. We only expect a small contribution of RBF measurements to this error, since RBF estimations are in very good agreement with Doppler OCT studies and have previously been shown to strongly correlate with in vivo blood flow metrics across a large BP range, as assessed by Laser Speckle Flowgraphy.^24,46,69-73^ However, in the previous subsection, we discussed how the more substantial error introduced by variability in oximetry measurements could have affected certain results. A lot of this variability is likely of technical nature, due to the documented presence of unaccounted non-uniform magnification, distortion, and illumination in particular parts of the retinal image, which can affect measurements, even with changes in the angle of gaze.^38,74^ These observations can explain the unexpected between-eye ODR differences that were present in certain retinal quadrants (Table S1). Of course, physiological effects could also result in additional optical artifacts.^75^ That said, due to incorporation of this expected variability to our original power analysis, our study was still able to show significant effects, despite the noise introduced by measurement uncertainty.

Secondly, this cross-sectional study cannot conclude if and how these initial BP-related alterations in O_2_ transport pertain to the development of ophthalmic pathologies. Due to the cross-sectional nature of the study, we also did not have robust information regarding the onset and duration of AHT. However, the incorporation of BP measurements from multiple previous occasions in the group definitions, as well as the requirement of uninterrupted use of antihypertensive medication over (at least) the past year resulted in almost all diagnoses occurring before at least three years.

Lastly, this population was almost entirely Caucasian and, as such, any results derived from it should not be immediately generalized to other ethnicities.

### Implications and conclusions

By showing that BP-related alterations in O_2_ transport may already exist in subjects with no signs of ophthalmic pathology, this study enhances our understanding of the baseline interplay between BP, RBF, and retinal oxygenation. This is important, but is merely a first step towards answering pertinent questions regarding the disease process. If hypoxia plays a role in complicated pathologies such as AMD and DR, when does the retina really become hypoxic and how early can we detect it in clinical practice?^7^ Can we use this information to evaluate treatments and slow disease progression? A special mention should be made at this point for glaucoma: according to the “chicken-egg” dilemma, structural loss could be both cause and consequence of impaired blood flow.^76^ In this regard, future studies could examine whether deficits in perfusion and oxygenation related to known glaucoma risk factors, such as BP status, can be helpful in stratifying the risk of incidence and/or progression of the disease.

In conclusion, we reported alterations in retinal oxygen delivery and extraction in ophthalmologically healthy subjects with different BP status. Future studies should incorporate the vascular supply of the choroid to elucidate whether increased oxygen extraction in uncontrolled AHT could be the result of compensatory mechanisms in effect. Longitudinal studies could investigate whether compromised delivery and extraction in subjects with low BP and treated AHT can explain the increased risk of glaucomatous damage in these population groups.

## Data Availability

All data is available upon reasonable request to the corresponding author.

