## Supplementary Table S1 for "Retinal oxygen delivery and extraction in ophthalmologically healthy subjects with different blood pressure status"

**Supplementary Table S1.** Regression coefficients of confounding factors associated with optical density ratios in multivariable analysis (only significant effects are included).

|  |  | **PI** | **Sphere** | **Cylinder (negative)** | **Disc area** | **Laterality**  **(0=OD, 1=OS)** |
| --- | --- | --- | --- | --- | --- | --- |
| **ST** | **ODR_art_** | -0.158 | *NA* | *NA* | *NA* | *NA* |
|  | **ODR_ven_** | -0.159 | *NA* | *NA* | *NA* | *NA* |
| **IT** | **ODR_art_** | *NA* | 0.016 | *NA* | *NA* | -0.102 |
|  | **ODR_ven_** | *NA* | *NA* | -0.062 | *NA* | -0.093 |
| **SN** | **ODR_art_** | *NA* | *NA* | *NA* | *NA* | *NA* |
|  | **ODR_ven_** | *NA* | *NA* | *NA* | *NA* | -0.096 |
| **IN** | **ODR_art_** | *NA* | *NA* | *NA* | +0.064 | +0.085 |
|  | **ODR_ven_** | -0.139 | *NA* | *NA* | *NA* | *NA* |
| ODR_art_, arterial optical density ratio; ODR_ven_, venous optical density ratio; ST, superotemporal; IT, inferotemporal; SN, superonasal; IN, inferonasal; PI, pigmentation index; OD, oculus dexter; OS, oculus sinister; *NA*, not applicable. | | | | | | |
