## Supplementary Table S2 for "Retinal oxygen delivery and extraction in ophthalmologically healthy subjects with different blood pressure status"

**Supplementary Table S2.** Corrected optical density ratios per retinal quadrant.

|  | **Arterial ODR_cor_** | **Venous ODR_cor_** | ***P* value** |
| --- | --- | --- | --- |
| **Global [mean (SD)]** | 0.138 (0.052) | 0.262 (0.052) | **2.5**$\boldsymbol{\cdot}$**10^-37^** |
| **ST [mean (SD)]** | 0.178 (0.081) | 0.269 (0.071) | **2.6**$\boldsymbol{\cdot}$**10^-19^** |
| **IT [mean (SD)]** | 0.170 (0.103) | 0.313 (0.109) | **3.2**$\boldsymbol{\cdot}$**10^-20^** |
| **SN [mean (SD)]** | 0.078 (0.115) | 0.202 (0.097) | **2.0**$\boldsymbol{\cdot}$**10^-16^** |
| **IN [mean (SD)]** | 0.124 (0.118) | 0.265 (0.086) | **3.7**$\boldsymbol{\cdot}$**10^-14^** |
| ***P* value** | **1.7**$\boldsymbol{\cdot}$**10^-10^** | **1.4**$\boldsymbol{\cdot}$**10^-13^** |  |
| ODR_cor_, corrected optical density ratio; ST, superotemporal; IT, inferotemporal; SN, superonasal; IN, inferonasal; SD, standard deviation. | | | |
